# Prevalence and Risks factors of the different forms of Domestic Violence Among Pregnant Women Attending Antenatal Clinics in the Buea Health District of Cameroon

**DOI:** 10.1101/2025.02.25.25322852

**Authors:** Elvis Asangbeng Tanue, Henrietta Nchang Chifen, Abdel Jélil Njouendou, Stanley Sunjo Nyuydzedze, Mildred Nje Laban, Dickson Shey Nsagha

## Abstract

**Background:** Domestic violence during pregnancy is a global public health problem which is linked with adverse maternal and fetal outcomes. Despite its significant impact on maternal and child health, the issue remains underexplored and underreported in Cameroon. This study therefore aimed at assess the various forms of domestic violence, their prevalences and associated factors among pregnant women attending antenatal care services in public and private hospitals in the Buea Health District of Cameroon.

**Materials and Methods:** This was a hospital-based cross-sectional study conducted between the months of April to June 2021. A consecutive sampling technique was used to select the participants and a, WHO instrument on domestic violence in low-income countries was used to assess violence against pregnant women. Data collected was cleaned and analysed using SPSS version 26.0. Multiple logistic regression analysis was used to determine the relationship between the dependent variable (domestic violence) and independent variables at a 95% confidence interval (CI) for types of domestic violence while adjusting for confounding factors.

**Results:** Of the 346 pregnant women who participated in this study, 303 (87.57%) were identified as experiencing at least one form of domestic violence. The most common form of domestic violence was psychological aggression (82.7%), followed by controlling behavior (78.0%), then physical (42.2%), economic deprivation (42.2%) and sexual (34.4%) violence. Women with monthly income below 50,000 XAF (AOR=4.31, 95% CI 1.23-15.08; *p=*0.022), and consuming alcohol (AOR=6.00, 95% CI 2.41-14.93; *p<*0.001) were more likely to experience psychological violence. Physical violence was associated with women with less than tertiary education (AOR=8.26, 95% CI 2.84-24.00; *p*=0.001) and having partners who gamble (AOR=8.11, 95% CI 3.34-19.69; *p*=0.001). Controlling behavior, was associated with partners who consumed alcohol (AOR 4.31; 95% CI 1.94-9.60; *p*=0.001) and to women with a monthly income below 25,000 XAF (AOR=3.53, 95% CI 1.26-9.89; *p*=0.016). Partners chosen by family (AOR=5.62, 95% CI 1.58-19.95; *p*=0.008), being housewives (AOR=9.55, 95% CI 2.36-38.57; *p*=0.002) and partners who smoked (AOR=3.22, 95% CI 1.32-7.84; p=0.010) were significantly associated to economic deprivation. Women who choose their partners (AOR=0.09, 95% CI 0.02-0.52; *p*=0.007) were less likely to experience sexual violence.

**Conclusion:** More than three quarters of the pregnant women in the study experience at least one type of domestic violence. This high proportion highlights the need for policy to address the problem of violence against pregnant women.

## 1.1 Background for the study

Violence against women is a global health and human right concern as it affects millions of women all over the world across ethnicity, culture, socioeconomic and educational class. [1]. Domestic violence or family violence against women from their intimate partners or other family members is prevalent across the world and has adverse implications on health especially during pregnancy where numerous health problems have been reported for both mothers and infants such as premature labour, low birth weight, emotional problems, mental health problems or even deaths in some cases. [2, 3]. According to a World Health Organization 2013 comprehensive review, ‘the global prevalence of physical and/or sexual, intimate partner violence among all ever-partnered women was estimated at 30%. The prevalence estimate ranged from 23.2% in high income countries to 24.6% in WHO Western pacific region. However, in the WHO African region, Eastern Mediterranean and South-East Asia regions, approximately 37% of ever-partnered women reportedly had experienced physical and/or sexual intimate partner violence at some point in their lives [4]. In sub-Saharan Africa, a study conducted by Kapiga, S et al, 2017 reported the prevalence of domestic violence at 61% [5]. A systematic review using 58 studies conducted by Muluneh, M. *et al*, 2020 which segregated the different forms of domestic violence reported 29.4% of emotional, 25.87% of physical and 18.87% of sexual violence [5]. The prevalence of domestic violence equally varies with regions in Africa, sub regional analysis found that women residing in the East 25% and West 30% African regions have experienced higher levels of emotional violence [6]. Recent data from the global database on violence against women in Cameroon gives life time prevalence of 51% for both physical and sexual partner violence. In the South West Region of Cameroon, more than 85% of people reported that women and girls experience rape, sexual violence and intimate partner violence [7].

Domestic violence has many different definitions, domestic violence is violence that takes place within a household and can be between any two people (parent and a child, siblings etc) within that household. It is however different from intimate partner violence (IPV) in that IPV occurs between romantic partners who may or may not be living together [8]. The United Nations High Commissioner for Human Rights defines violence against women in article 1 of its declaration as ‘‘any act of gender-based violence that results in, or is likely to result in, physical, sexual or psychological harm or suffering to women, including threats of such acts, coercion or arbitrary deprivation of liberty, whether occurring in public or private life [9]. Domestic violence can also be seen as a pattern of abusive behavior in any relationship that is used by someone to gain or maintain control over another in a close relationship [10]. Domestic violence looks beyond violence perpetrated by a male or partner in an intimate relationship to violence carried out by any member of the family on another in a family relationship. Domestic violence against women encompasses any physical, sexual or emotional, financial, verbal, controlling behavior abuses imposed upon women in family relationships [11]. During pregnancy, violence against women in pregnancy doesn’t only affect the mother but the fetus as well [12,13]. Domestic violence and maternal mortality ratios in low-income countries are recognized as a global public health problem with serious consequences on both maternal and fetal health [13, 14,15]. In high income countries the problem is still widespread, Greece for example has a prevalence of 6% with 3.4% experiencing violence mainly by their husbands or partners since the beginning of pregnancy. [16]. The rates of domestic violence in Africa remain high. Kenya according to a study by Luhumyo, L, 2020 had a prevalence of 34.4% of women who have at least witnessed one form of violence during pregnancy [17]. Across Ethiopia, the prevalence ranges from 20.6% to 59.0% within north to south Ethiopia [18, 19, 20, 21] and this high prevalence are being associated with cultural, socio demographic and personal characteristics. In Cameroon, it is estimated from the Demographic Health Survey (DHS) data in 2004 that approximately 38.7% of women experienced physical violence, 30.7% emotional violence and 14.8% sexual violence from intimate partners throughout their lifetime. [22]. The demographic survey aggregated data for all women, it is therefore essential to determine the magnitude of the problem among pregnant women as well as establishing key predictors to facilitate control and management as well as policy development.

Domestic violence during pregnancy has both fatal and non-fatal adverse health outcomes for the pregnant woman and her baby raging from direct, physiological effects of stress to physical and mental health effects on fetal growth and development [21].

Violence during pregnancy negatively affects the quality of life of women after delivery, impairing both physical and mental health [23]. Physical injuries like bruises, abrasions, lacerations, broken bones, attempted strangulations, [23], psychological and emotional suffering lead to behavioral changes such as alcoholism, recreational drug abuse, smoking, post-traumatic stress disorder, unsafe sexual behaviors and poor self-esteem [24]. Malayoto, L., et al (2013) found strong association between women who experience physical, sexual and psychological violence during pregnancy and mental health disorders including sleep disorders, anxiety and depression [25]. Pregnant women experiencing violence are 3 times more likely to have prenatal deaths than those who do not undergo violence [12].

The risk factors of DV are a combination of societal, community, relational and individual factors. In Bangladesh, violence against women is deeply rooted in patriarchy laws where it is considered legitimate right of the husband [28]. Several studies on risk factors for domestic violence in pregnancy reported; traditional gender norms, poverty, unemployment, lack of education of both partners, having experiences of violence before pregnancy, alcohol, tobacco use, age of marriage, unplanned pregnancy, late initiation of ANC, partners use of alcohol, disobedience of women towards husbands, husband with history of arrest, rural residency [20,21, 46].

Despite the heavy consequences of domestic violence, it is usually not reported by the women because of stigma, fear of losing their children, feeling of shame, denial, improper judgment and lack of personal resources to either leave the home or change the situation according to studies conducted by Shamu s, et al, 2011 and Omolo, J & Kamweya, A. M. 2014[33,34].

Armed conflict has significant impact on both individuals and families living in conflict affected areas, Devakumar et al, 2021, identified 8 qualitative studies which showed men’s experience of conflict including financial stresses contributes to perpetration of family violence. [3]. There remains a gap in knowledge on the prevalence and determinants of family violence (domestic violence) in conflict affected areas such as the case in South West region of Cameroon, We therefore aimed to explore the prevalence and predictors of different forms of domestic violence among pregnant women attending Antenatal care clinic services in Buea Health District. The findings from this study are expected to provide information on the magnitude of domestic violence among pregnant women in the South West region and key factors associated with the different forms identified among pregnant women attending ANC in the South West region important to support policy on early prevention, identification and care.

## Materials and Methods

### Study area and setting

The study was carried out in the Buea Health District, in the South West region of Cameroon. It is the capital city of the South West region which is one of the regions affected by the anglophone crises in Cameroon. This was a hospital-based study which targeted pregnant women visiting or attending antenatal care services in 4 health areas in the Buea Health District. Data collection took place between the months of May and June 2021.

A structured questionnaire adapted from WHO tool for assessing domestic violence in low-income countries [35,36] was used for data collection. The adapted questionnaire was pre-tested at Bokwango health center and necessary modifications to the questionnaire were made to ensure the information collected was relevant.

#### 3.5 Sample size determination for the cross-sectional study

The sample size for this study was calculated using the Lorentz Formula.

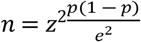

Where; Z= 1.96 at 95% confidence level.

p= prevalence of domestic violence to be 33% [75].

e= 5% or 0.05; which is the level of precision

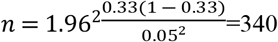

A total of 346 participants were recruited for the quantitative part of this study.

### Sampling technique

Four health areas were randomly selected from the 7 health areas in Buea health district. Due to the ongoing crises in the region, some facilities were not functioning, all functional health facilities (private and public) in the health areas selected were enrolled. Proportionate sampling was used to determine the number of participants per health area. Convenient sampling was used to recruit participants from among the pregnant women attending ANC in health facilities during ANC days.

#### Inclusion Criteria

Pregnant women attending ANC services in selected health facility in Buea health district.

#### Exclusion criteria

Pregnant woman or women admitted or critically ill.

### Data Management and Statistical Analysis

The data collected was verified, entered in to Microsoft excel 2013 and analysed using SPSS version 25.0. All data was kept confidential in password protected folder and computer. The questionnaires had no information that could identify participants such as names or telephone numbers. All filled questionnaires were locked in a cupboard and were coded. Exploratory analysis was done to check missing values and outliers. Descriptive statistics were used to obtain frequencies, percentages and means. Bivariate logistic regression analysis was used to determine the association between each of the independent variables and occurrence of domestic violence (outcome variable). Thereafter, independent variables with *p* < 0.20 in bivariate logistic regression analysis were included in the multivariate logistic regression model to adjust for possible confounders. The statistical significance was set at *p*<0.05.

### Ethical considerations

Ethical clearance was obtained from the Faculty of Health Sciences Institutional Review Board of the University of Buea (Reference number: 2021/1369-04/UB/SG/IRB/FHS). Administrative approval was obtained from the Regional Delegation of the Ministry of Public Health for the South West Region and the administration of the health facilities where the study was carried out. Before data collection, the purpose of the study was explained to participants and their guardians. Written informed consent was obtained from each participant and the guardians of participants under 18 years old. After receiving guardian’s consent, written assent was also obtained from the participants themselves. The questionnaires were coded to ensure confidentiality.

## Results

A total of 346 pregnant women were recruited from health facilities in 4 health areas of the Buea Health District. The mean age of the participants was 26.8 ± 5.3 years. Majority of the women (59%) were married, with 90.6% of the ever-married women in monogamous marriages. Table 1a shows the demographic characteristics of the study participants.

**Table 1a:**
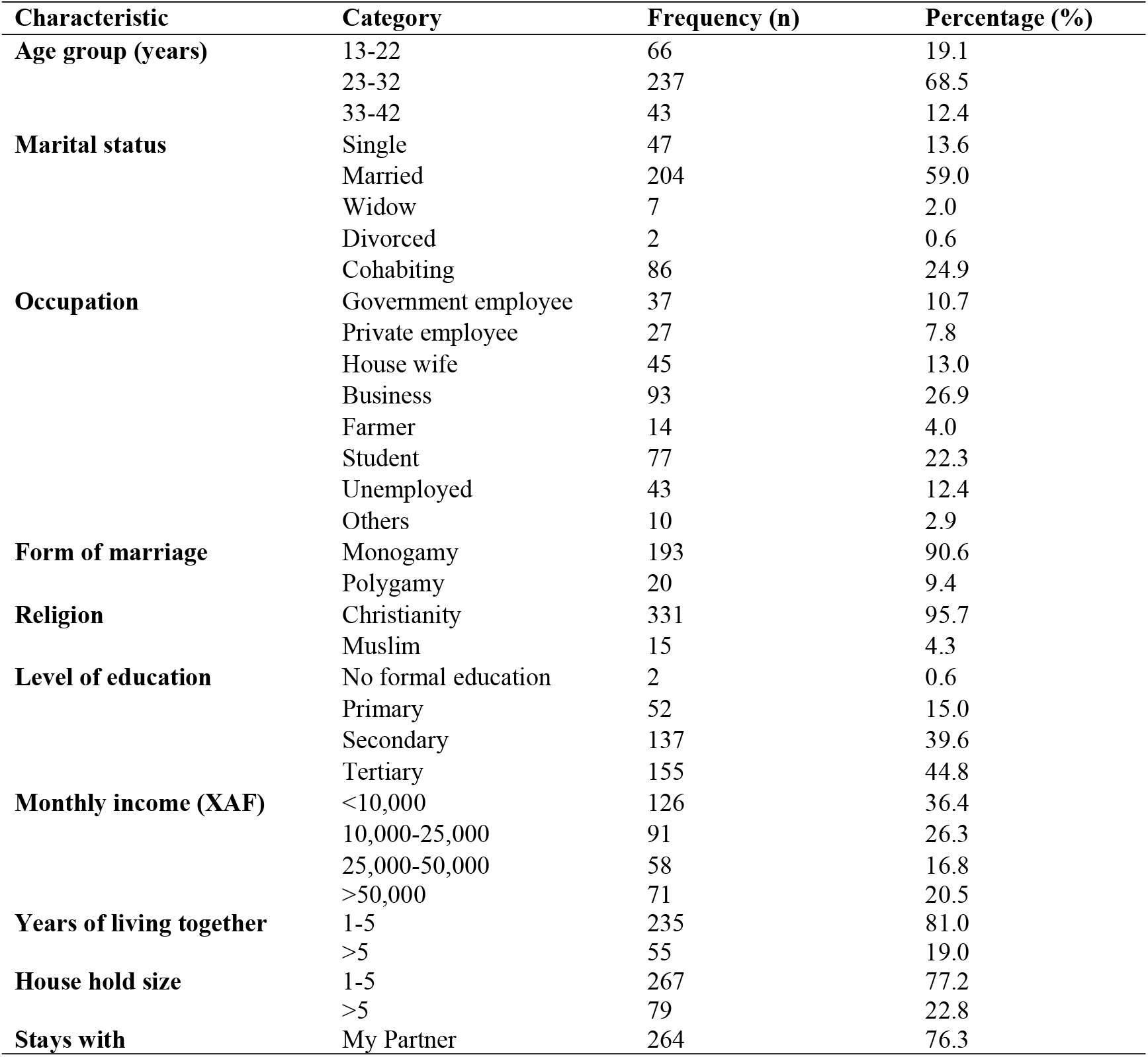
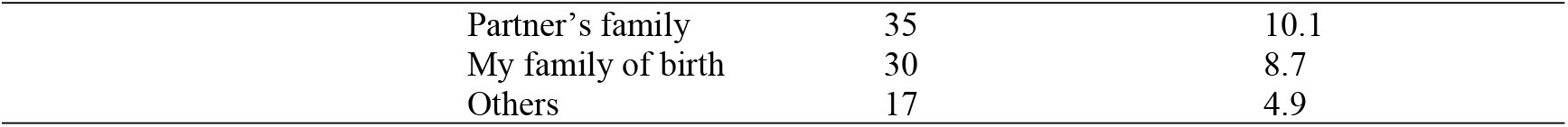
Socio-demographic Characteristics of Pregnant Women in Buea Health District.

### Prevalence of domestic violence in Buea health district

Of the 346 participants recruited, 303 (87.57%) had experienced at least one form of domestic violence. Some of the participants experienced a combination of 2 or more types of violence, 27.7% of the participants experience atleast 2 types of violence, 19.9% 3 types, 12.4%, 4 types and 19.1% atleast 5 types of violence. Figure 1. shows the prevalence of the various types of domestic among pregnant women in the Buea health district.

**Figure 1:**
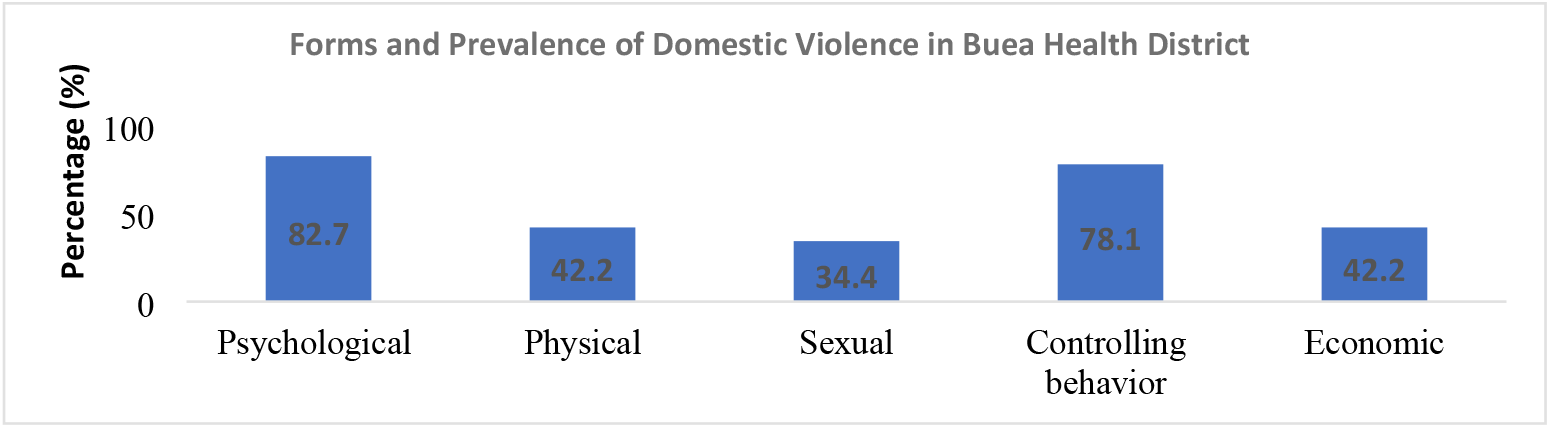
Prevalence of Types of Domestic Violence in Buea Health District.

### Determinants of Psychological Violence during pregnancy

Women who received low monthly income of 10,000-25,000 XAF were about 4 times more likely to experience psychological violence compared to those who received greater than 50,000 XAF (AOR 4.31; 95% CI 1.23-15.08; *p* = 0.022). Women who consume alcohol were 6 times more likely to experience psychological violence than those who do not (AOR 6.00; 95% CI 2.41-14.93; *p* < 0.001). Similarly pregnant women whose partners consume alcohol were more than 3 times more likely to experience psychological violence compared to those whose partners do not consume alcohol (AOR 3.93; 95% CI 1.57-9.81; *p* = 0.003) (Table 2).

**Table 2:**
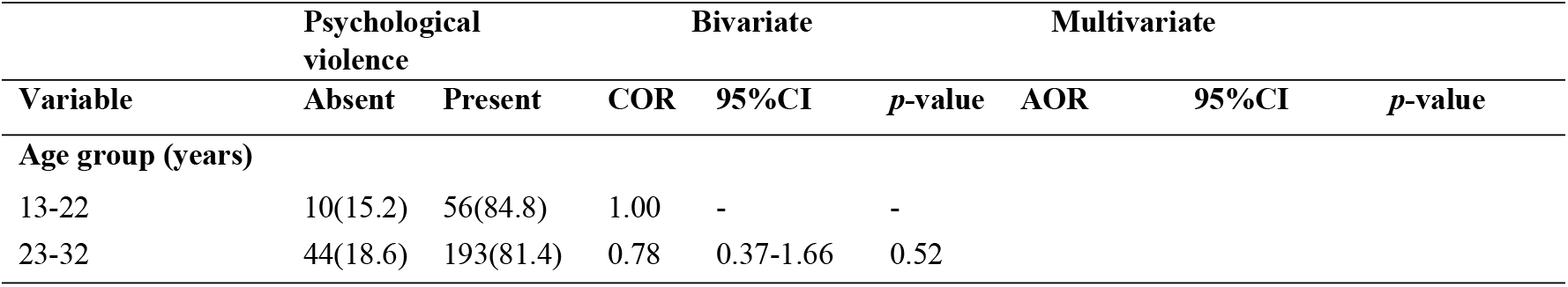
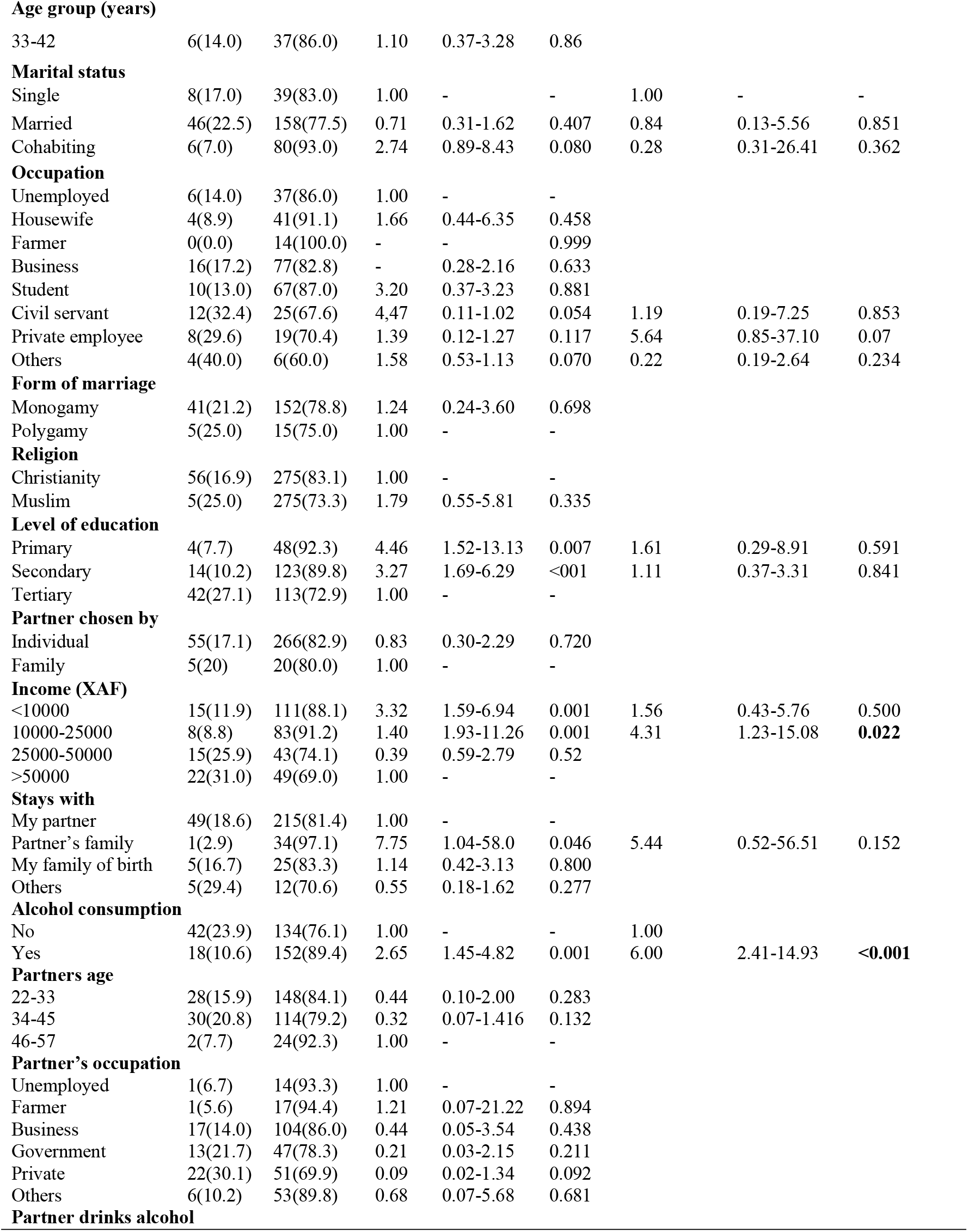
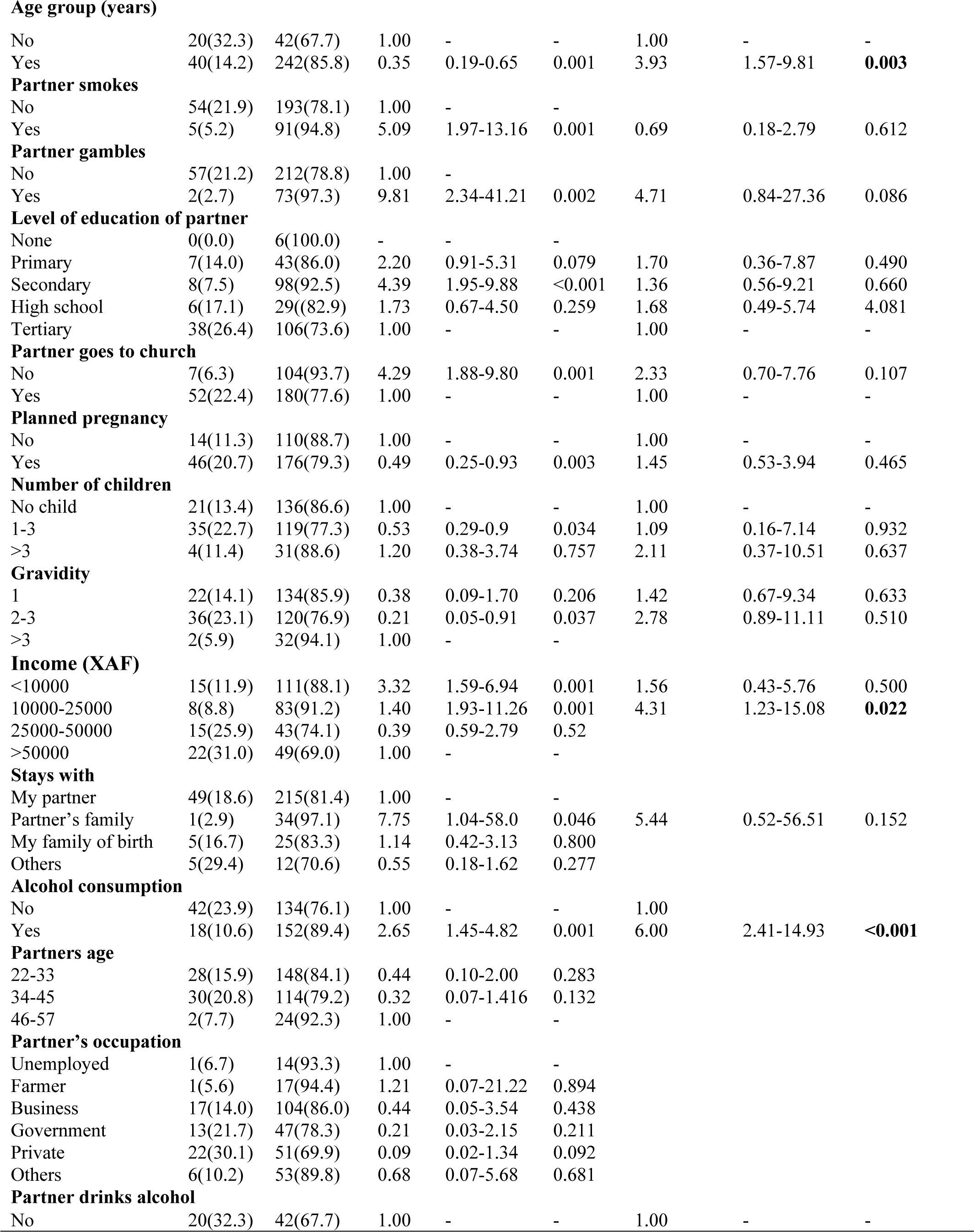
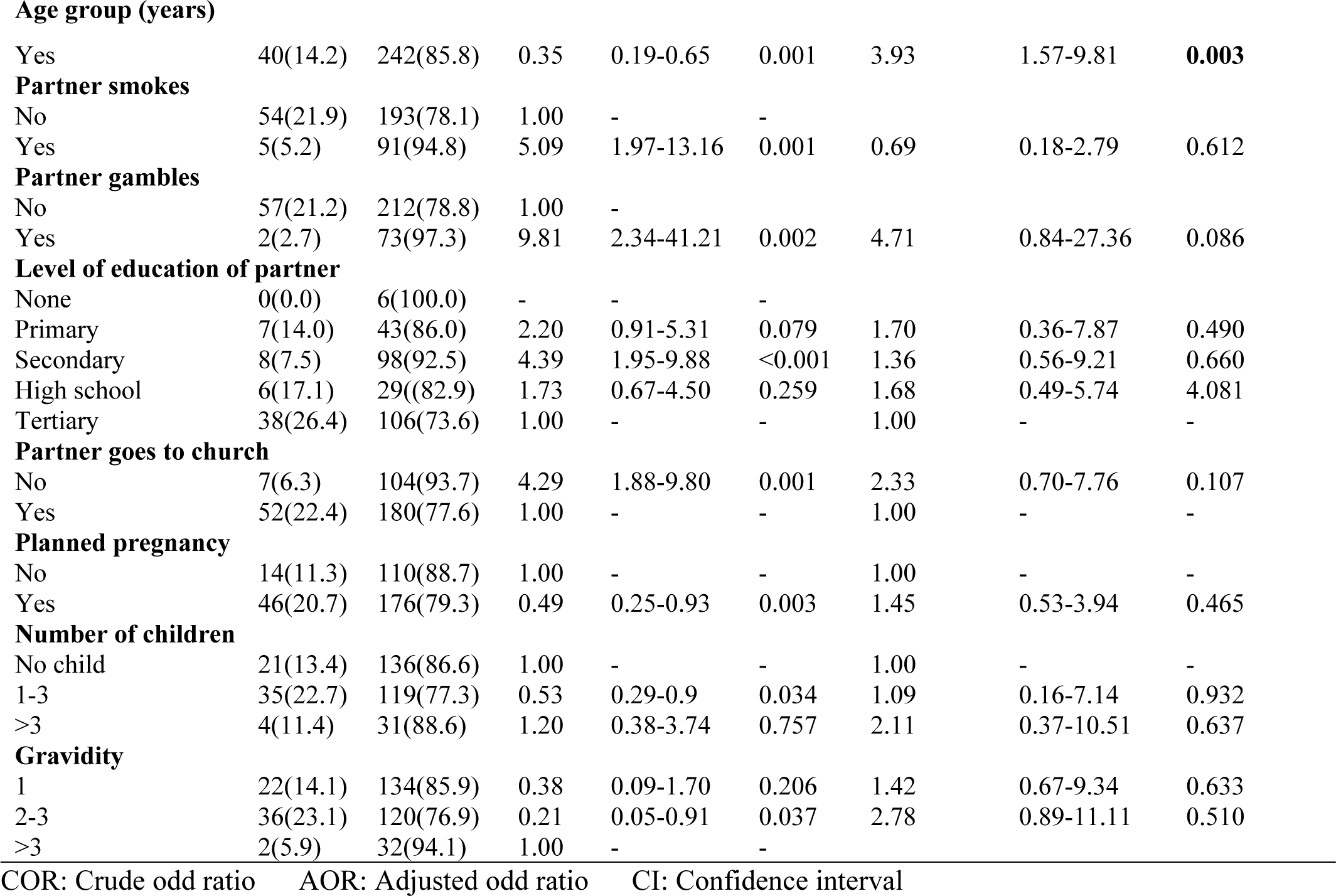
Determinants of Psychological Violence during pregnancy.

| Variable | Psychological violence |  | Bivariate |  |  | Multivariate |  |  |
| --- | --- | --- | --- | --- | --- | --- | --- | --- |
|  | Absent | Present | COR | 95%CI | *p*-value | AOR | 95%CI | *p*-value |
| **Age group (years)** |  |  |  |  |  |  |  |  |
| 13-22 | 10(15.2) | 56(84.8) | 1.00 | - | - |  |  |  |
| 23-32 | 44(18.6) | 193(81.4) | 0.78 | 0.37-1.66 | 0.52 |  |  |  |

|  | **Psychological violence** |  | **Bivariate** |  |  | **Multivariate** |  |  |
| --- | --- | --- | --- | --- | --- | --- | --- | --- |
| **Variable** | **Absent** | **Present** | **COR** | **95%CI** | **p-value** | **AOR** | **95%CI** | **p-value** |
| **Age group (years)** |  |  |  |  |  |  |  |  |
| 33-42 | 6(14.0) | 37(86.0) | 1.10 | 0.37-3.28 | 0.86 |  |  |  |
| **Marital status** |  |  |  |  |  |  |  |  |
| Single | 8(17.0) | 39(83.0) | 1.00 | - | - | 1.00 | - | - |
| Married | 46(22.5) | 158(77.5) | 0.71 | 0.31-1.62 | 0.407 | 0.84 | 0.13-5.56 | 0.851 |
| Cohabiting | 6(7.0) | 80(93.0) | 2.74 | 0.89-8.43 | 0.080 | 0.28 | 0.31-26.41 | 0.362 |
| **Occupation** |  |  |  |  |  |  |  |  |
| Unemployed | 6(14.0) | 37(86.0) | 1.00 | - | - |  |  |  |
| Housewife | 4(8.9) | 41(91.1) | 1.66 | 0.44-6.35 | 0.458 |  |  |  |
| Farmer | 0(0.0) | 14(100.0) | - | - | 0.999 |  |  |  |
| Business | 16(17.2) | 77(82.8) | - | 0.28-2.16 | 0.633 |  |  |  |
| Student | 10(13.0) | 67(87.0) | 3.20 | 0.37-3.23 | 0.881 |  |  |  |
| Civil servant | 12(32.4) | 25(67.6) | 4.47 | 0.11-1.02 | 0.054 | 1.19 | 0.19-7.25 | 0.853 |
| Private employee | 8(29.6) | 19(70.4) | 1.39 | 0.12-1.27 | 0.117 | 5.64 | 0.85-37.10 | 0.07 |
| Others | 4(40.0) | 6(60.0) | 1.58 | 0.53-1.13 | 0.070 | 0.22 | 0.19-2.64 | 0.234 |
| **Form of marriage** |  |  |  |  |  |  |  |  |
| Monogamy | 41(21.2) | 152(78.8) | 1.24 | 0.24-3.60 | 0.698 |  |  |  |
| Polygamy | 5(25.0) | 15(75.0) | 1.00 | - | - |  |  |  |
| **Religion** |  |  |  |  |  |  |  |  |
| Christianity | 56(16.9) | 275(83.1) | 1.00 | - | - |  |  |  |
| Muslim | 5(25.0) | 275(73.3) | 1.79 | 0.55-5.81 | 0.335 |  |  |  |
| **Level of education** |  |  |  |  |  |  |  |  |
| Primary | 4(7.7) | 48(92.3) | 4.46 | 1.52-13.13 | 0.007 | 1.61 | 0.29-8.91 | 0.591 |
| Secondary | 14(10.2) | 123(89.8) | 3.27 | 1.69-6.29 | <001 | 1.11 | 0.37-3.31 | 0.841 |
| Tertiary | 42(27.1) | 113(72.9) | 1.00 | - | - |  |  |  |
| **Partner chosen by** |  |  |  |  |  |  |  |  |
| Individual | 55(17.1) | 266(82.9) | 0.83 | 0.30-2.29 | 0.720 |  |  |  |
| Family | 5(20) | 20(80.0) | 1.00 | - | - |  |  |  |
| **Income (XAF)** |  |  |  |  |  |  |  |  |
| <10000 | 15(11.9) | 111(88.1) | 3.32 | 1.59-6.94 | 0.001 | 1.56 | 0.43-5.76 | 0.500 |
| 10000-25000 | 8(8.8) | 83(91.2) | 1.40 | 1.93-11.26 | 0.001 | 4.31 | 1.23-15.08 | **0.022** |
| 25000-50000 | 15(25.9) | 43(74.1) | 0.39 | 0.59-2.79 | 0.52 |  |  |  |
| >50000 | 22(31.0) | 49(69.0) | 1.00 | - | - |  |  |  |
| **Stays with** |  |  |  |  |  |  |  |  |
| My partner | 49(18.6) | 215(81.4) | 1.00 | - | - |  |  |  |
| Partner's family | 1(2.9) | 34(97.1) | 7.75 | 1.04-58.0 | 0.046 | 5.44 | 0.52-56.51 | 0.152 |
| My family of birth | 5(16.7) | 25(83.3) | 1.14 | 0.42-3.13 | 0.800 |  |  |  |
| Others | 5(29.4) | 12(70.6) | 0.55 | 0.18-1.62 | 0.277 |  |  |  |
| **Alcohol consumption** |  |  |  |  |  |  |  |  |
| No | 42(23.9) | 134(76.1) | 1.00 | - | - | 1.00 |  |  |
| Yes | 18(10.6) | 152(89.4) | 2.65 | 1.45-4.82 | 0.001 | 6.00 | 2.41-14.93 | **<0.001** |
| **Partners age** |  |  |  |  |  |  |  |  |
| 22-33 | 28(15.9) | 148(84.1) | 0.44 | 0.10-2.00 | 0.283 |  |  |  |
| 34-45 | 30(20.8) | 114(79.2) | 0.32 | 0.07-1.416 | 0.132 |  |  |  |
| 46-57 | 2(7.7) | 24(92.3) | 1.00 | - | - |  |  |  |
| **Partner's occupation** |  |  |  |  |  |  |  |  |
| Unemployed | 1(6.7) | 14(93.3) | 1.00 | - | - |  |  |  |
| Farmer | 1(5.6) | 17(94.4) | 1.21 | 0.07-21.22 | 0.894 |  |  |  |
| Business | 17(14.0) | 104(86.0) | 0.44 | 0.05-3.54 | 0.438 |  |  |  |
| Government | 13(21.7) | 47(78.3) | 0.21 | 0.03-2.15 | 0.211 |  |  |  |
| Private | 22(30.1) | 51(69.9) | 0.09 | 0.02-1.34 | 0.092 |  |  |  |
| Others | 6(10.2) | 53(89.8) | 0.68 | 0.07-5.68 | 0.681 |  |  |  |
| **Partner drinks alcohol** |  |  |  |  |  |  |  |  |

|  | Psychological violence |  | Bivariate |  |  | Multivariate |  |  |
| --- | --- | --- | --- | --- | --- | --- | --- | --- |
| Variable | Absent | Present | COR | 95%CI | p-value | AOR | 95%CI | p-value |
| **Age group (years)** |  |  |  |  |  |  |  |  |
| No | 20(32.3) | 42(67.7) | 1.00 | - | - | 1.00 | - | - |
| Yes | 40(14.2) | 242(85.8) | 0.35 | 0.19-0.65 | 0.001 | 3.93 | 1.57-9.81 | **0.003** |
| **Partner smokes** |  |  |  |  |  |  |  |  |
| No | 54(21.9) | 193(78.1) | 1.00 | - | - |  |  |  |
| Yes | 5(5.2) | 91(94.8) | 5.09 | 1.97-13.16 | 0.001 | 0.69 | 0.18-2.79 | 0.612 |
| **Partner gambles** |  |  |  |  |  |  |  |  |
| No | 57(21.2) | 212(78.8) | 1.00 | - | - |  |  |  |
| Yes | 2(2.7) | 73(97.3) | 9.81 | 2.34-41.21 | 0.002 | 4.71 | 0.84-27.36 | 0.086 |
| **Level of education of partner** |  |  |  |  |  |  |  |  |
| None | 0(0.0) | 6(100.0) | - | - | - |  |  |  |
| Primary | 7(14.0) | 43(86.0) | 2.20 | 0.91-5.31 | 0.079 | 1.70 | 0.36-7.87 | 0.490 |
| Secondary | 8(7.5) | 98(92.5) | 4.39 | 1.95-9.88 | <0.001 | 1.36 | 0.56-9.21 | 0.660 |
| High school | 6(17.1) | 29(82.9) | 1.73 | 0.67-4.50 | 0.259 | 1.68 | 0.49-5.74 | 4.081 |
| Tertiary | 38(26.4) | 106(73.6) | 1.00 | - | - | 1.00 | - | - |
| **Partner goes to church** |  |  |  |  |  |  |  |  |
| No | 7(6.3) | 104(93.7) | 4.29 | 1.88-9.80 | 0.001 | 2.33 | 0.70-7.76 | 0.107 |
| Yes | 52(22.4) | 180(77.6) | 1.00 | - | - | 1.00 | - | - |
| **Planned pregnancy** |  |  |  |  |  |  |  |  |
| No | 14(11.3) | 110(88.7) | 1.00 | - | - | 1.00 | - | - |
| Yes | 46(20.7) | 176(79.3) | 0.49 | 0.25-0.93 | 0.003 | 1.45 | 0.53-3.94 | 0.465 |
| **Number of children** |  |  |  |  |  |  |  |  |
| No child | 21(13.4) | 136(86.6) | 1.00 | - | - | 1.00 | - | - |
| 1-3 | 35(22.7) | 119(77.3) | 0.53 | 0.29-0.9 | 0.034 | 1.09 | 0.16-7.14 | 0.932 |
| >3 | 4(11.4) | 31(88.6) | 1.20 | 0.38-3.74 | 0.757 | 2.11 | 0.37-10.51 | 0.637 |
| **Gravidity** |  |  |  |  |  |  |  |  |
| 1 | 22(14.1) | 134(85.9) | 0.38 | 0.09-1.70 | 0.206 | 1.42 | 0.67-9.34 | 0.633 |
| 2-3 | 36(23.1) | 120(76.9) | 0.21 | 0.05-0.91 | 0.037 | 2.78 | 0.89-11.11 | 0.510 |
| >3 | 2(5.9) | 32(94.1) | 1.00 | - | - |  |  |  |
| **Income (XAF)** |  |  |  |  |  |  |  |  |
| <10000 | 15(11.9) | 111(88.1) | 3.32 | 1.59-6.94 | 0.001 | 1.56 | 0.43-5.76 | 0.500 |
| 10000-25000 | 8(8.8) | 83(91.2) | 1.40 | 1.93-11.26 | 0.001 | 4.31 | 1.23-15.08 | **0.022** |
| 25000-50000 | 15(25.9) | 43(74.1) | 0.39 | 0.59-2.79 | 0.52 |  |  |  |
| >50000 | 22(31.0) | 49(69.0) | 1.00 | - | - |  |  |  |
| **Stays with** |  |  |  |  |  |  |  |  |
| My partner | 49(18.6) | 215(81.4) | 1.00 | - | - |  |  |  |
| Partner's family | 1(2.9) | 34(97.1) | 7.75 | 1.04-58.0 | 0.046 | 5.44 | 0.52-56.51 | 0.152 |
| My family of birth | 5(16.7) | 25(83.3) | 1.14 | 0.42-3.13 | 0.800 |  |  |  |
| Others | 5(29.4) | 12(70.6) | 0.55 | 0.18-1.62 | 0.277 |  |  |  |
| **Alcohol consumption** |  |  |  |  |  |  |  |  |
| No | 42(23.9) | 134(76.1) | 1.00 | - | - | 1.00 |  |  |
| Yes | 18(10.6) | 152(89.4) | 2.65 | 1.45-4.82 | 0.001 | 6.00 | 2.41-14.93 | **<0.001** |
| **Partners age** |  |  |  |  |  |  |  |  |
| 22-33 | 28(15.9) | 148(84.1) | 0.44 | 0.10-2.00 | 0.283 |  |  |  |
| 34-45 | 30(20.8) | 114(79.2) | 0.32 | 0.07-1.416 | 0.132 |  |  |  |
| 46-57 | 2(7.7) | 24(92.3) | 1.00 | - | - |  |  |  |
| **Partner's occupation** |  |  |  |  |  |  |  |  |
| Unemployed | 1(6.7) | 14(93.3) | 1.00 | - | - |  |  |  |
| Farmer | 1(5.6) | 17(94.4) | 1.21 | 0.07-21.22 | 0.894 |  |  |  |
| Business | 17(14.0) | 104(86.0) | 0.44 | 0.05-3.54 | 0.438 |  |  |  |
| Government | 13(21.7) | 47(78.3) | 0.21 | 0.03-2.15 | 0.211 |  |  |  |
| Private | 22(30.1) | 51(69.9) | 0.09 | 0.02-1.34 | 0.092 |  |  |  |
| Others | 6(10.2) | 53(89.8) | 0.68 | 0.07-5.68 | 0.681 |  |  |  |
| **Partner drinks alcohol** |  |  |  |  |  |  |  |  |
| No | 20(32.3) | 42(67.7) | 1.00 | - | - | 1.00 | - | - |
|  | Absent | Present | COR | 95%CI | p-value | AOR | 95%CI | p-value |
| **Age group (years)** |  |  |  |  |  |  |  |  |
| Yes | 40(14.2) | 242(85.8) | 0.35 | 0.19-0.65 | 0.001 | 3.93 | 1.57-9.81 | **0.003** |
| **Partner smokes** |  |  |  |  |  |  |  |  |
| No | 54(21.9) | 193(78.1) | 1.00 | - | - |  |  |  |
| Yes | 5(5.2) | 91(94.8) | 5.09 | 1.97-13.16 | 0.001 | 0.69 | 0.18-2.79 | 0.612 |
| **Partner gambles** |  |  |  |  |  |  |  |  |
| No | 57(21.2) | 212(78.8) | 1.00 | - | - |  |  |  |
| Yes | 2(2.7) | 73(97.3) | 9.81 | 2.34-41.21 | 0.002 | 4.71 | 0.84-27.36 | 0.086 |
| **Level of education of partner** |  |  |  |  |  |  |  |  |
| None | 0(0.0) | 6(100.0) | - | - | - |  |  |  |
| Primary | 7(14.0) | 43(86.0) | 2.20 | 0.91-5.31 | 0.079 | 1.70 | 0.36-7.87 | 0.490 |
| Secondary | 8(7.5) | 98(92.5) | 4.39 | 1.95-9.88 | <0.001 | 1.36 | 0.56-9.21 | 0.660 |
| High school | 6(17.1) | 29(82.9) | 1.73 | 0.67-4.50 | 0.259 | 1.68 | 0.49-5.74 | 4.081 |
| Tertiary | 38(26.4) | 106(73.6) | 1.00 | - | - | 1.00 | - | - |
| **Partner goes to church** |  |  |  |  |  |  |  |  |
| No | 7(6.3) | 104(93.7) | 4.29 | 1.88-9.80 | 0.001 | 2.33 | 0.70-7.76 | 0.107 |
| Yes | 52(22.4) | 180(77.6) | 1.00 | - | - | 1.00 | - | - |
| **Planned pregnancy** |  |  |  |  |  |  |  |  |
| No | 14(11.3) | 110(88.7) | 1.00 | - | - | 1.00 | - | - |
| Yes | 46(20.7) | 176(79.3) | 0.49 | 0.25-0.93 | 0.003 | 1.45 | 0.53-3.94 | 0.465 |
| **Number of children** |  |  |  |  |  |  |  |  |
| No child | 21(13.4) | 136(86.6) | 1.00 | - | - | 1.00 | - | - |
| 1-3 | 35(22.7) | 119(77.3) | 0.53 | 0.29-0.9 | 0.034 | 1.09 | 0.16-7.14 | 0.932 |
| >3 | 4(11.4) | 31(88.6) | 1.20 | 0.38-3.74 | 0.757 | 2.11 | 0.37-10.51 | 0.637 |
| **Gravidity** |  |  |  |  |  |  |  |  |
| 1 | 22(14.1) | 134(85.9) | 0.38 | 0.09-1.70 | 0.206 | 1.42 | 0.67-9.34 | 0.633 |
| 2-3 | 36(23.1) | 120(76.9) | 0.21 | 0.05-0.91 | 0.037 | 2.78 | 0.89-11.11 | 0.510 |
| >3 | 2(5.9) | 32(94.1) | 1.00 | - | - |  |  |  |

COR: Crude odd ratio AOR: Adjusted odd ratio CI: Confidence interval

### Determinants of Physical violence during pregnancy

Women with primary education or less and women who had secondary education, were about 8 times and 3 times respectively more likely to suffer physical violence than women with tertiary education (AOR 8.26; 95% CI 2.84-24.00; *p* = 0.001), (AOR 3.42; 95%V CI I.65-7.08; *p* = 0.001). Those who lived in households greater than 5 persons were more than 2 times more likely to experience physical violence compared to those in households of at most 5 persons (AOR 2.86;95% CI 1.02-8.03; *p* = 0.003). Women whose partners aged between 46-57 years had more than 2 times negative association towards physical violence than women who had partners who were much younger ie between 22-33 years. Women whose partners smoke were greater than 2 times more likely to experience physical violence than women whose partners do not smoke (AOR 2.83;95% CI 1.31-6.14; *p* = 0.008) and women whose partners gamble had about 8 times more odds of experiencing physical violence than women whose partners neither smoke nor gamble (AOR 8.11 95% CI 3.34-19.69; *p* = 0.001) (Table 5).

**Table 5:**
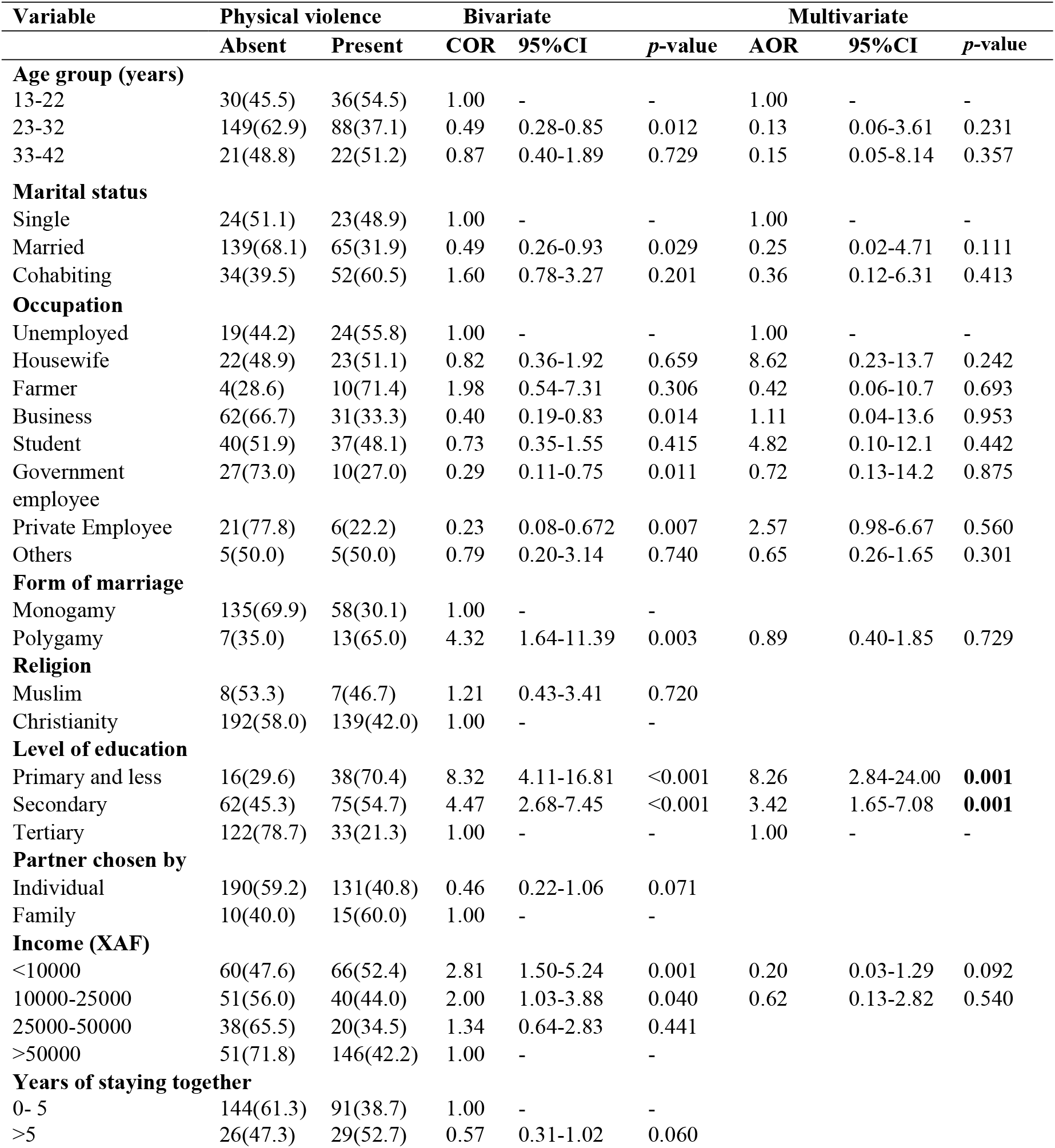
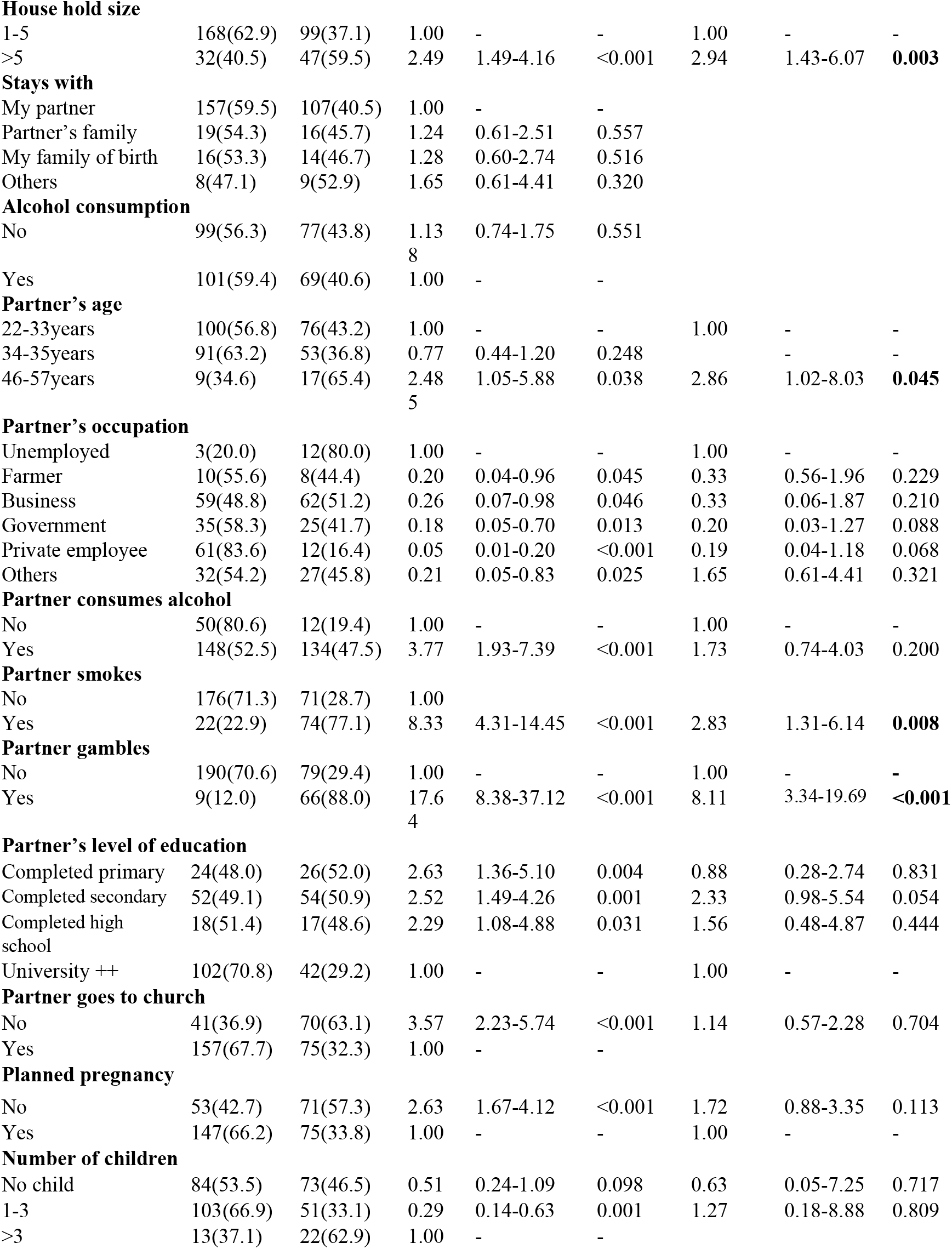
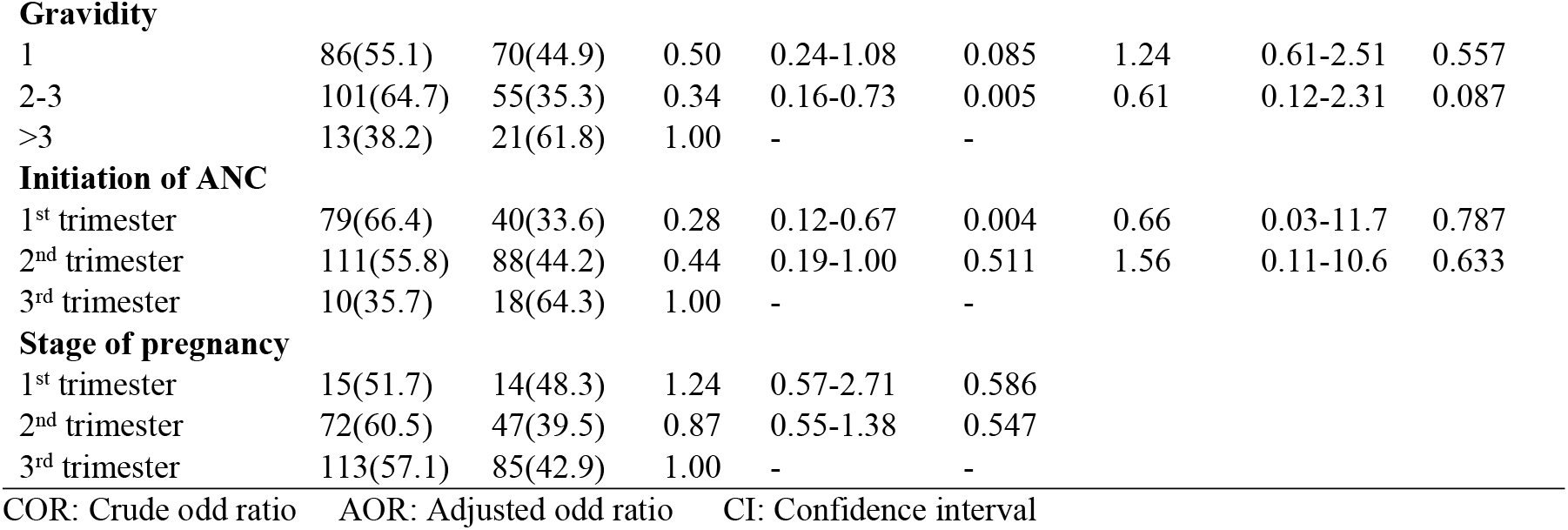
Determinants of Physical violence during pregnancy.

Table 6 shows the results of the multivariate analysis between the sociodemographic characteristics of participants, partner’s characteristics and obstetric characteristics of participants. Participants who chose their partners had a 91% less likelihood to be sexually violated by their partners compared to those whose partners were chosen by family members (AOR 0.09; 95% CI 0.02-0.52; *p* = 0.007). Women whose partners were 34-45 years and 46-57 years had respectively 76% and 73% less chances to experience sexual violence than women whose partners were 22-33 years (AOR 0.24;95% CI 0.07-0.72; *p* = 0.010) and (AOR 0.27; 95% CI 0.09-0.77; *p* = 0.015). Women whose partners gamble had about 3 times more odds of experiencing sexual violence than those whose partners do not gamble (AOR 3.24; 95% CI 1.54-44.18; *p* = 0.014).

**Table 6:**
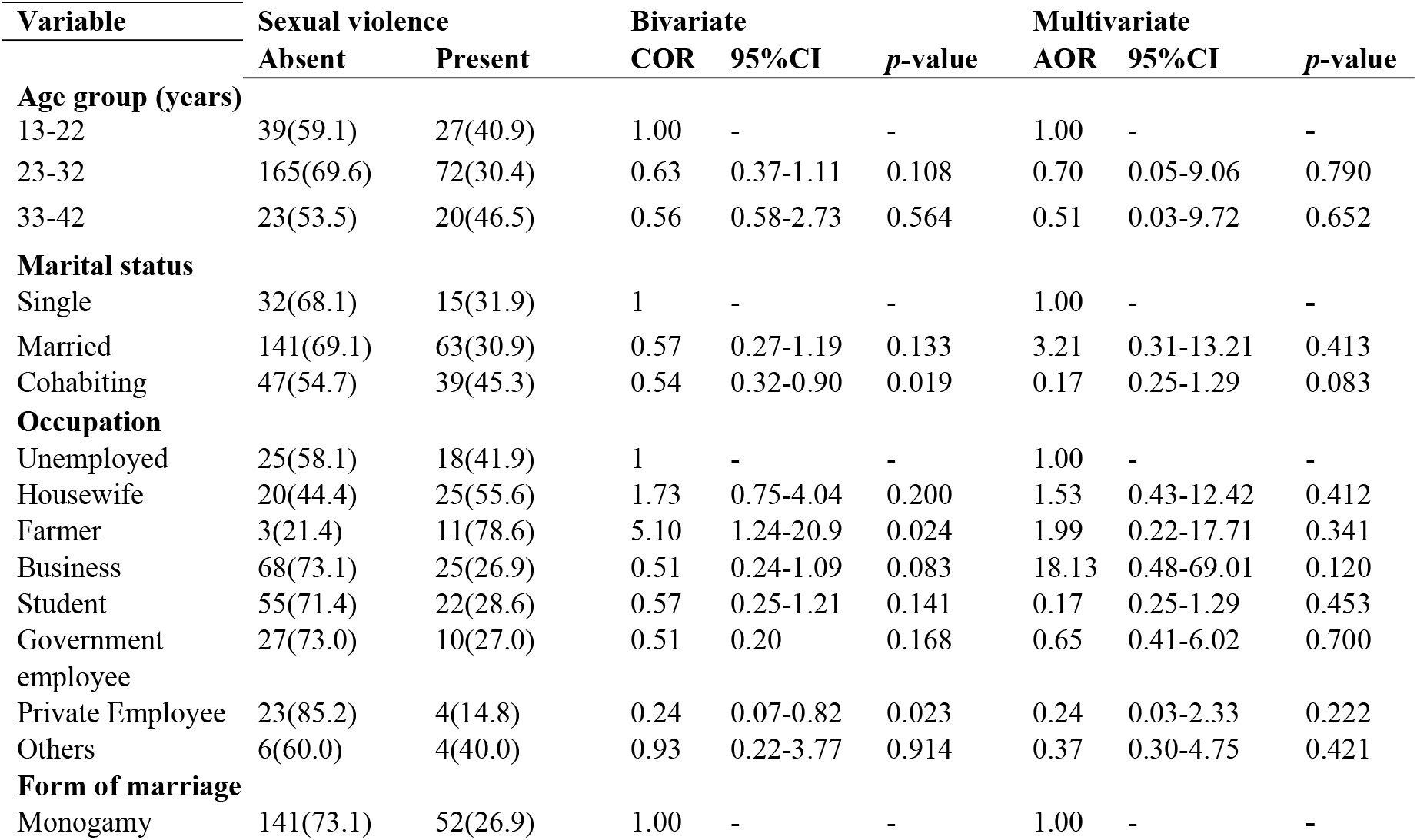
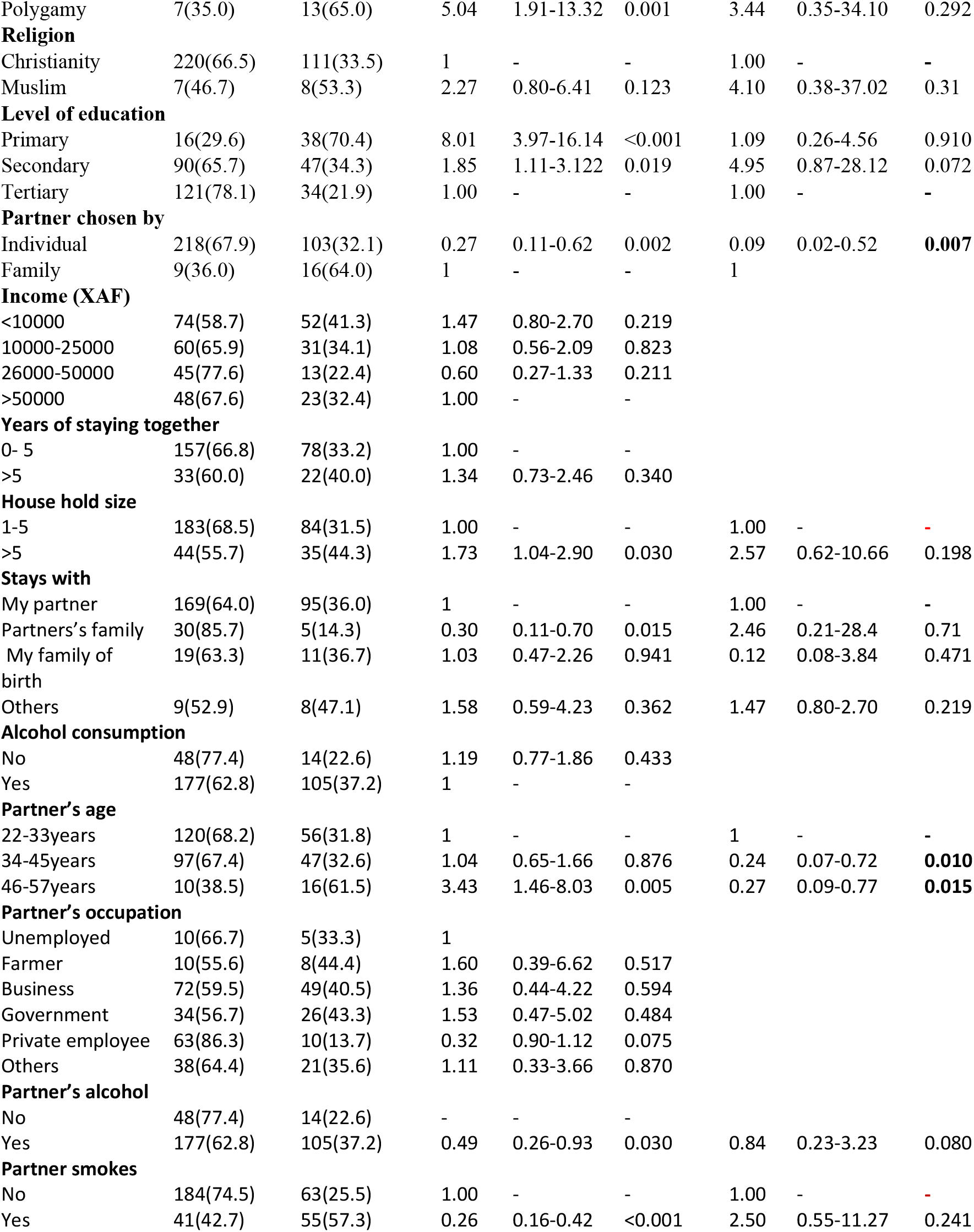
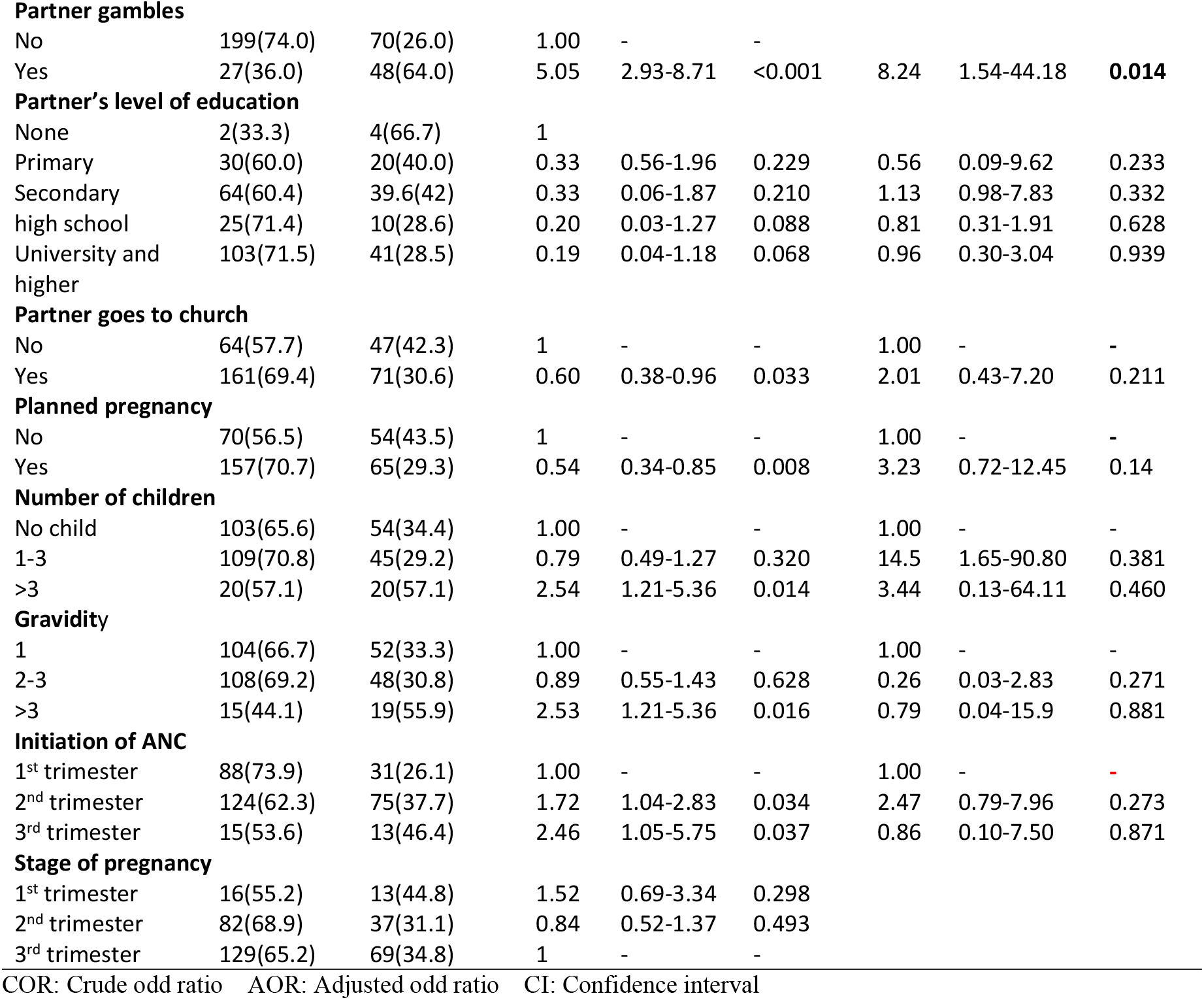
Determinants of Sexual violence during pregnancy.

| Variable | Sexual violence |  | Bivariate |  |  | Multivariate |  |  |
| --- | --- | --- | --- | --- | --- | --- | --- | --- |
|  | Absent | Present | COR | 95%CI | p-value | AOR | 95%CI | p-value |
| **Age group (years)** |  |  |  |  |  |  |  |  |
| 13-22 | 39(59.1) | 27(40.9) | 1.00 | - | - | 1.00 | - | - |
| 23-32 | 165(69.6) | 72(30.4) | 0.63 | 0.37-1.11 | 0.108 | 0.70 | 0.05-9.06 | 0.790 |
| 33-42 | 23(53.5) | 20(46.5) | 0.56 | 0.58-2.73 | 0.564 | 0.51 | 0.03-9.72 | 0.652 |
| **Marital status** |  |  |  |  |  |  |  |  |
| Single | 32(68.1) | 15(31.9) | 1 | - | - | 1.00 | - | - |
| Married | 141(69.1) | 63(30.9) | 0.57 | 0.27-1.19 | 0.133 | 3.21 | 0.31-13.21 | 0.413 |
| Cohabiting | 47(54.7) | 39(45.3) | 0.54 | 0.32-0.90 | 0.019 | 0.17 | 0.25-1.29 | 0.083 |
| **Occupation** |  |  |  |  |  |  |  |  |
| Unemployed | 25(58.1) | 18(41.9) | 1 | - | - | 1.00 | - | - |
| Housewife | 20(44.4) | 25(55.6) | 1.73 | 0.75-4.04 | 0.200 | 1.53 | 0.43-12.42 | 0.412 |
| Farmer | 3(21.4) | 11(78.6) | 5.10 | 1.24-20.9 | 0.024 | 1.99 | 0.22-17.71 | 0.341 |
| Business | 68(73.1) | 25(26.9) | 0.51 | 0.24-1.09 | 0.083 | 18.13 | 0.48-69.01 | 0.120 |
| Student | 55(71.4) | 22(28.6) | 0.57 | 0.25-1.21 | 0.141 | 0.17 | 0.25-1.29 | 0.453 |
| Government employee | 27(73.0) | 10(27.0) | 0.51 | 0.20 | 0.168 | 0.65 | 0.41-6.02 | 0.700 |
| Private Employee | 23(85.2) | 4(14.8) | 0.24 | 0.07-0.82 | 0.023 | 0.24 | 0.03-2.33 | 0.222 |
| Others | 6(60.0) | 4(40.0) | 0.93 | 0.22-3.77 | 0.914 | 0.37 | 0.30-4.75 | 0.421 |
| **Form of marriage** |  |  |  |  |  |  |  |  |
| Monogamy | 141(73.1) | 52(26.9) | 1.00 | - | - | 1.00 | - | - |
|  | Absent | Present | COR | 95%CI | p-value | AOR | 95%CI | p-value |
| Polygamy | 7(35.0) | 13(65.0) | 5.04 | 1.91-13.32 | 0.001 | 3.44 | 0.35-34.10 | 0.292 |
| **Religion** |  |  |  |  |  |  |  |  |
| Christianity | 220(66.5) | 111(33.5) | 1 | - | - | 1.00 | - | - |
| Muslim | 7(46.7) | 8(53.3) | 2.27 | 0.80-6.41 | 0.123 | 4.10 | 0.38-37.02 | 0.31 |
| **Level of education** |  |  |  |  |  |  |  |  |
| Primary | 16(29.6) | 38(70.4) | 8.01 | 3.97-16.14 | <0.001 | 1.09 | 0.26-4.56 | 0.910 |
| Secondary | 90(65.7) | 47(34.3) | 1.85 | 1.11-3.122 | 0.019 | 4.95 | 0.87-28.12 | 0.072 |
| Tertiary | 121(78.1) | 34(21.9) | 1.00 | - | - | 1.00 | - | - |
| **Partner chosen by** |  |  |  |  |  |  |  |  |
| Individual | 218(67.9) | 103(32.1) | 0.27 | 0.11-0.62 | 0.002 | 0.09 | 0.02-0.52 | **0.007** |
| Family | 9(36.0) | 16(64.0) | 1 | - | - | 1 |  |  |
| **Income (XAF)** |  |  |  |  |  |  |  |  |
| <10000 | 74(58.7) | 52(41.3) | 1.47 | 0.80-2.70 | 0.219 |  |  |  |
| 10000-25000 | 60(65.9) | 31(34.1) | 1.08 | 0.56-2.09 | 0.823 |  |  |  |
| 26000-50000 | 45(77.6) | 13(22.4) | 0.60 | 0.27-1.33 | 0.211 |  |  |  |
| >50000 | 48(67.6) | 23(32.4) | 1.00 | - | - |  |  |  |
| **Years of staying together** |  |  |  |  |  |  |  |  |
| 0- 5 | 157(66.8) | 78(33.2) | 1.00 | - | - |  |  |  |
| >5 | 33(60.0) | 22(40.0) | 1.34 | 0.73-2.46 | 0.340 |  |  |  |
| **House hold size** |  |  |  |  |  |  |  |  |
| 1-5 | 183(68.5) | 84(31.5) | 1.00 | - | - | 1.00 | - | - |
| >5 | 44(55.7) | 35(44.3) | 1.73 | 1.04-2.90 | 0.030 | 2.57 | 0.62-10.66 | 0.198 |
| **Stays with** |  |  |  |  |  |  |  |  |
| My partner | 169(64.0) | 95(36.0) | 1 | - | - | 1.00 | - | - |
| Partners's family | 30(85.7) | 5(14.3) | 0.30 | 0.11-0.70 | 0.015 | 2.46 | 0.21-28.4 | 0.71 |
| My family of birth | 19(63.3) | 11(36.7) | 1.03 | 0.47-2.26 | 0.941 | 0.12 | 0.08-3.84 | 0.471 |
| Others | 9(52.9) | 8(47.1) | 1.58 | 0.59-4.23 | 0.362 | 1.47 | 0.80-2.70 | 0.219 |
| **Alcohol consumption** |  |  |  |  |  |  |  |  |
| No | 48(77.4) | 14(22.6) | 1.19 | 0.77-1.86 | 0.433 |  |  |  |
| Yes | 177(62.8) | 105(37.2) | 1 | - | - |  |  |  |
| **Partner's age** |  |  |  |  |  |  |  |  |
| 22-33years | 120(68.2) | 56(31.8) | 1 | - | - | 1 | - | - |
| 34-45years | 97(67.4) | 47(32.6) | 1.04 | 0.65-1.66 | 0.876 | 0.24 | 0.07-0.72 | **0.010** |
| 46-57years | 10(38.5) | 16(61.5) | 3.43 | 1.46-8.03 | 0.005 | 0.27 | 0.09-0.77 | **0.015** |
| **Partner's occupation** |  |  |  |  |  |  |  |  |
| Unemployed | 10(66.7) | 5(33.3) | 1 |  |  |  |  |  |
| Farmer | 10(55.6) | 8(44.4) | 1.60 | 0.39-6.62 | 0.517 |  |  |  |
| Business | 72(59.5) | 49(40.5) | 1.36 | 0.44-4.22 | 0.594 |  |  |  |
| Government | 34(56.7) | 26(43.3) | 1.53 | 0.47-5.02 | 0.484 |  |  |  |
| Private employee | 63(86.3) | 10(13.7) | 0.32 | 0.90-1.12 | 0.075 |  |  |  |
| Others | 38(64.4) | 21(35.6) | 1.11 | 0.33-3.66 | 0.870 |  |  |  |
| **Partner's alcohol** |  |  |  |  |  |  |  |  |
| No | 48(77.4) | 14(22.6) | - | - | - |  |  |  |
| Yes | 177(62.8) | 105(37.2) | 0.49 | 0.26-0.93 | 0.030 | 0.84 | 0.23-3.23 | 0.080 |
| **Partner smokes** |  |  |  |  |  |  |  |  |
| No | 184(74.5) | 63(25.5) | 1.00 | - | - | 1.00 | - | - |
| Yes | 41(42.7) | 55(57.3) | 0.26 | 0.16-0.42 | <0.001 | 2.50 | 0.55-11.27 | 0.241 |
|  | Absent | Present | COR | 95%CI | p-value | AOR | 95%CI | p-value |
| **Partner gambles** |  |  |  |  |  |  |  |  |
| No | 199(74.0) | 70(26.0) | 1.00 | - | - |  |  |  |
| Yes | 27(36.0) | 48(64.0) | 5.05 | 2.93-8.71 | <0.001 | 8.24 | 1.54-44.18 | **0.014** |
| **Partner's level of education** |  |  |  |  |  |  |  |  |
| None | 2(33.3) | 4(66.7) | 1 |  |  |  |  |  |
| Primary | 30(60.0) | 20(40.0) | 0.33 | 0.56-1.96 | 0.229 | 0.56 | 0.09-9.62 | 0.233 |
| Secondary | 64(60.4) | 39.6(42) | 0.33 | 0.06-1.87 | 0.210 | 1.13 | 0.98-7.83 | 0.332 |
| high school | 25(71.4) | 10(28.6) | 0.20 | 0.03-1.27 | 0.088 | 0.81 | 0.31-1.91 | 0.628 |
| University and higher | 103(71.5) | 41(28.5) | 0.19 | 0.04-1.18 | 0.068 | 0.96 | 0.30-3.04 | 0.939 |
| **Partner goes to church** |  |  |  |  |  |  |  |  |
| No | 64(57.7) | 47(42.3) | 1 | - | - | 1.00 | - | - |
| Yes | 161(69.4) | 71(30.6) | 0.60 | 0.38-0.96 | 0.033 | 2.01 | 0.43-7.20 | 0.211 |
| **Planned pregnancy** |  |  |  |  |  |  |  |  |
| No | 70(56.5) | 54(43.5) | 1 | - | - | 1.00 | - | - |
| Yes | 157(70.7) | 65(29.3) | 0.54 | 0.34-0.85 | 0.008 | 3.23 | 0.72-12.45 | 0.14 |
| **Number of children** |  |  |  |  |  |  |  |  |
| No child | 103(65.6) | 54(34.4) | 1.00 | - | - | 1.00 | - | - |
| 1-3 | 109(70.8) | 45(29.2) | 0.79 | 0.49-1.27 | 0.320 | 14.5 | 1.65-90.80 | 0.381 |
| >3 | 20(57.1) | 20(57.1) | 2.54 | 1.21-5.36 | 0.014 | 3.44 | 0.13-64.11 | 0.460 |
| **Gravidity** |  |  |  |  |  |  |  |  |
| 1 | 104(66.7) | 52(33.3) | 1.00 | - | - | 1.00 | - | - |
| 2-3 | 108(69.2) | 48(30.8) | 0.89 | 0.55-1.43 | 0.628 | 0.26 | 0.03-2.83 | 0.271 |
| >3 | 15(44.1) | 19(55.9) | 2.53 | 1.21-5.36 | 0.016 | 0.79 | 0.04-15.9 | 0.881 |
| **Initiation of ANC** |  |  |  |  |  |  |  |  |
| 1<sup>st</sup> trimester | 88(73.9) | 31(26.1) | 1.00 | - | - | 1.00 | - | - |
| 2<sup>nd</sup> trimester | 124(62.3) | 75(37.7) | 1.72 | 1.04-2.83 | 0.034 | 2.47 | 0.79-7.96 | 0.273 |
| 3<sup>rd</sup> trimester | 15(53.6) | 13(46.4) | 2.46 | 1.05-5.75 | 0.037 | 0.86 | 0.10-7.50 | 0.871 |
| **Stage of pregnancy** |  |  |  |  |  |  |  |  |
| 1<sup>st</sup> trimester | 16(55.2) | 13(44.8) | 1.52 | 0.69-3.34 | 0.298 |  |  |  |
| 2<sup>nd</sup> trimester | 82(68.9) | 37(31.1) | 0.84 | 0.52-1.37 | 0.493 |  |  |  |
| 3<sup>rd</sup> trimester | 129(65.2) | 69(34.8) | 1 | - | - |  |  |  |

COR: Crude odd ratio AOR: Adjusted odd ratio CI: Confidence interval

#### 4.3.5 Determinants of Economic violence during pregnancy

Women who were house wives were more than 9 times more likely to experience economic violence compared to women who were unemployed (AOR=9.55, 95% CI 2.36-38.57; *P*-value=0.002). Women whose family chose partners for them had more than 5 times higher odds of experiencing economic violence than those who choose their partners themselves (AOR =5.62, 95% CI 1.58-19.95; *P*-value=0.008). Women whose partners smoked had 3 times higher the risk of experiencing economic violence than women whose partners do not smoke (AOR= 3.22, 95% CI 1.32-7.84; p-value=0.010). More so, women with partners who gamble were more than 4 times more likely to economic violence than women whose partners do not (AOR 4.79, 95% CI 2.05-11.19; *P*-value=<0.001). Finally, women who started antenatal at 2^nd^ trimester had about 2 times higher chances of experiencing economic violence than women who started at first trimester (AOR 2.26, 95% CI 1.03-4.95; *P*-value=0.041). Regression analysis results of economic violence are presented on table 7.

**Table 7:**
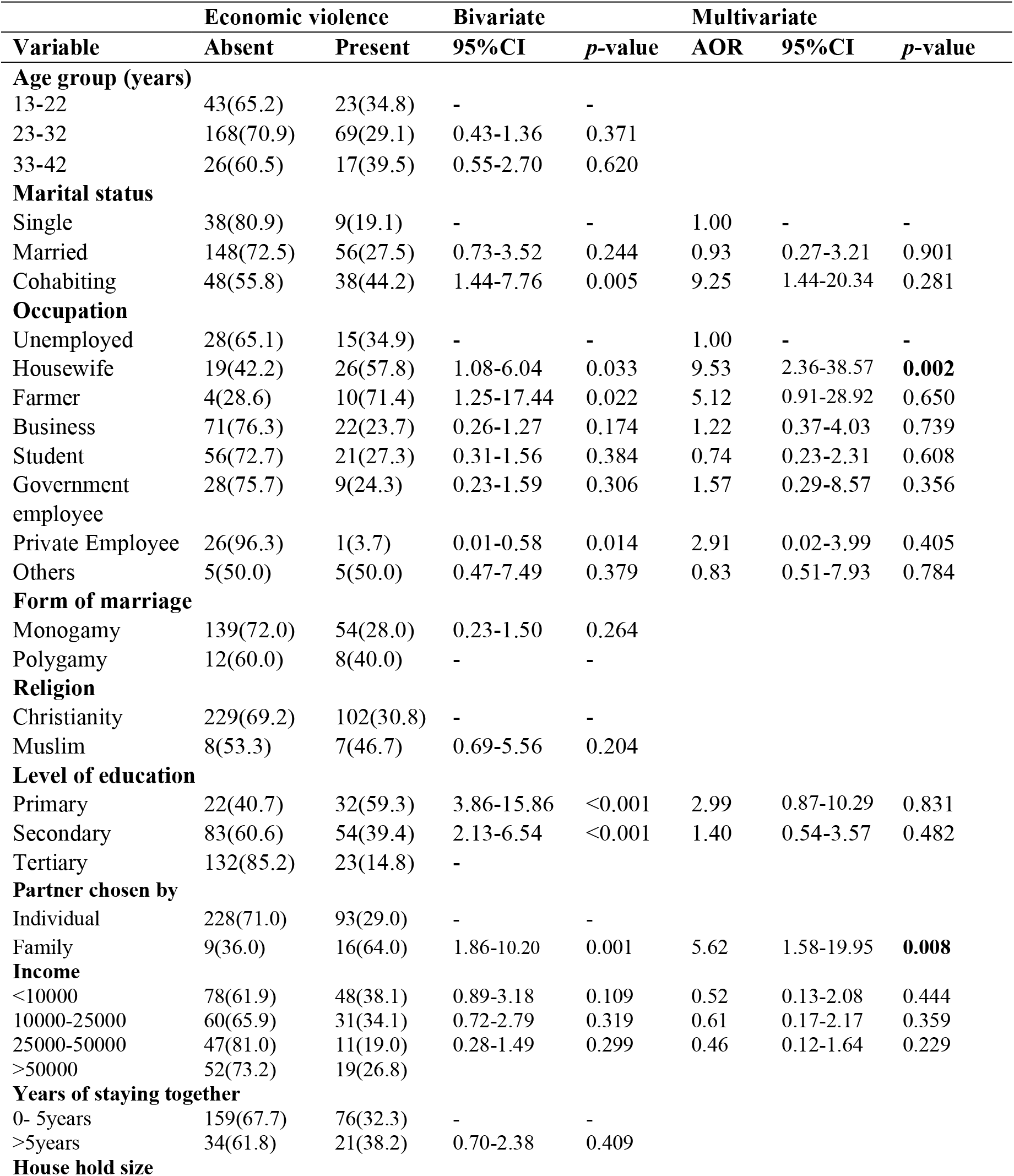
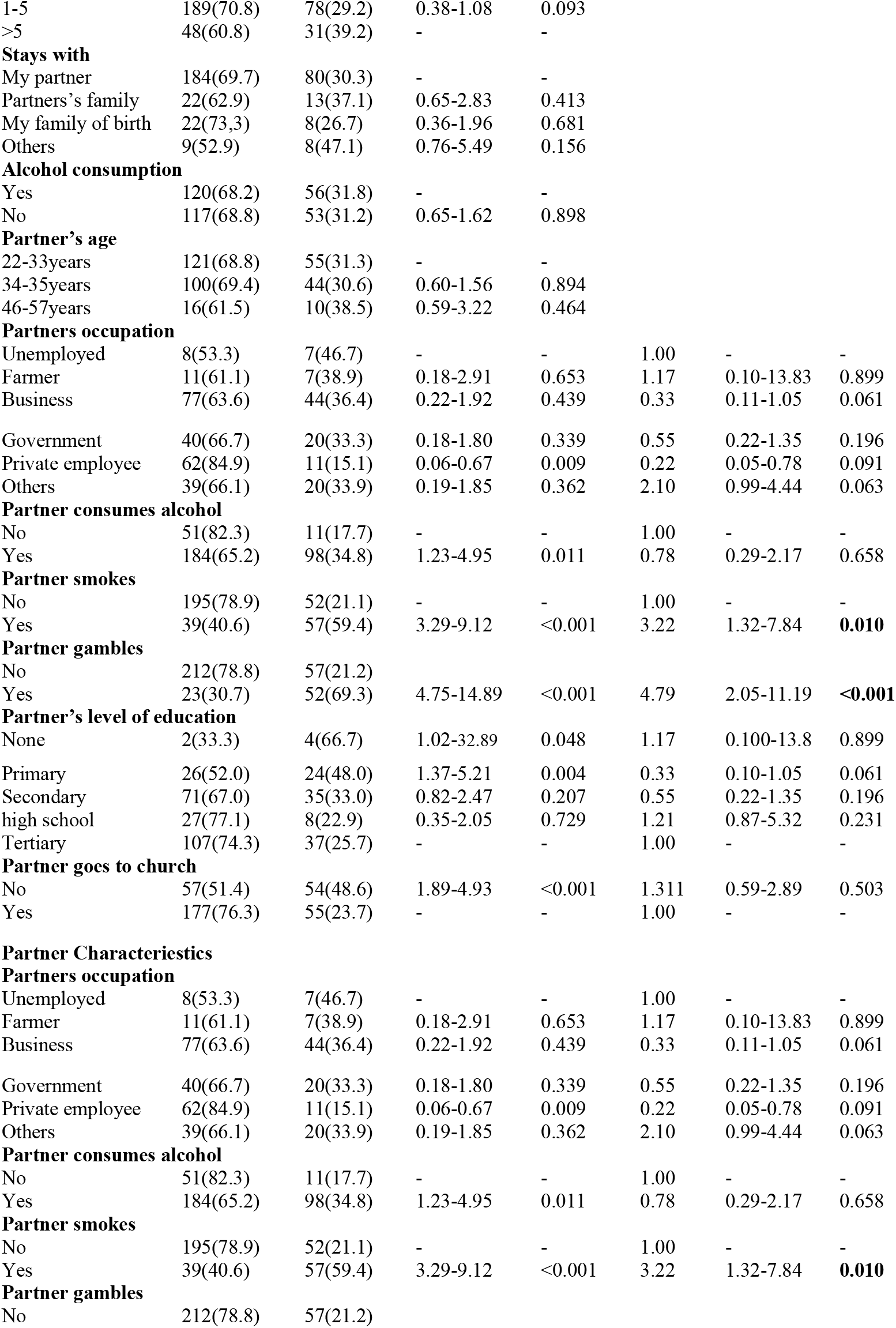
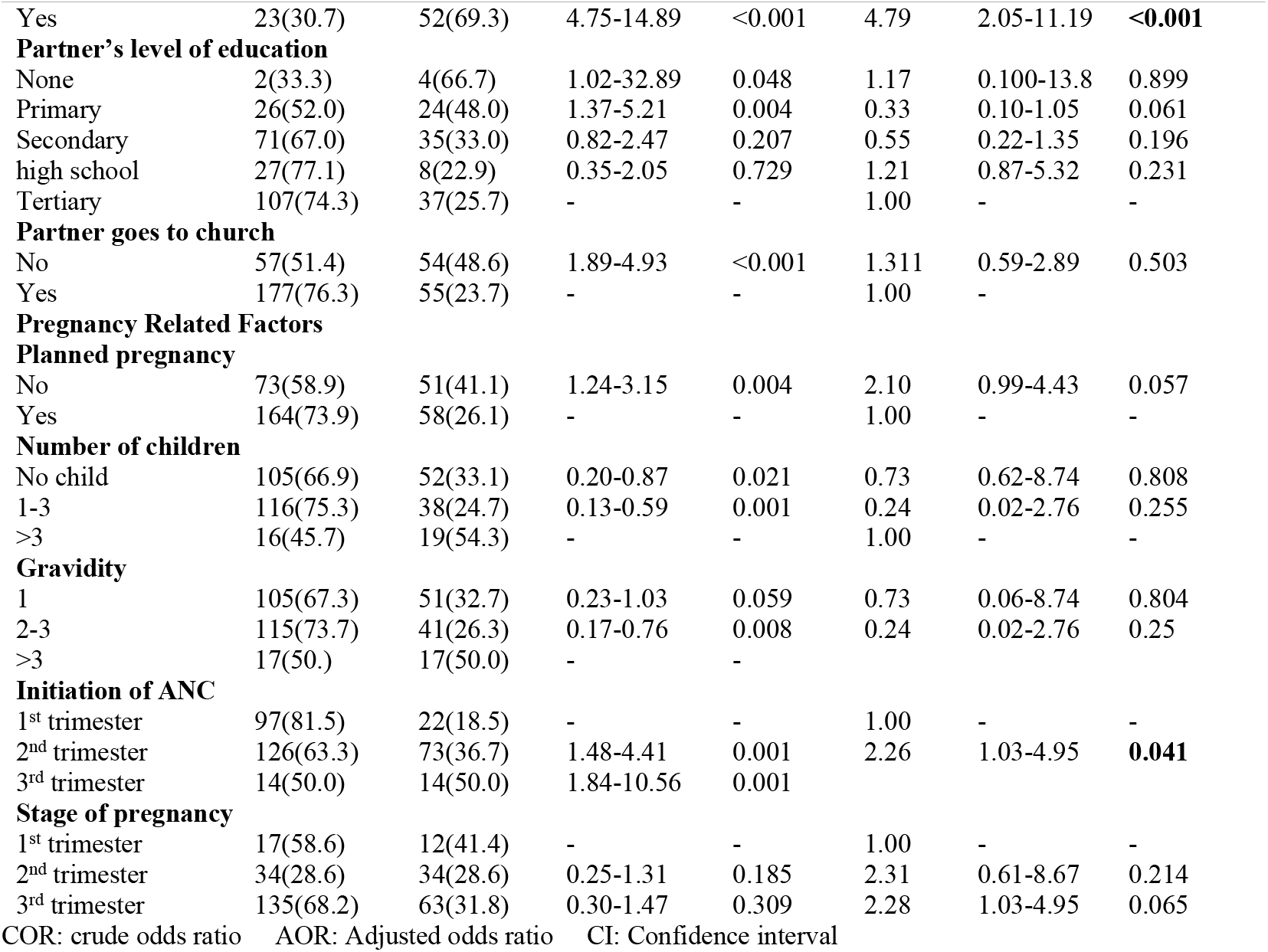
Determinants of Economic violence during pregnancy.

## Discussion

Domestic violence continues to prevail in all parts of the world predominating in low-income countries with life threatening and serious health consequences on both the woman and fetus. The overall prevalence of domestic violence during pregnancy in this study was 87.57%. The prevalence obtained in this study is similar to that of studies conducted in Bangladesh 87% [41]. However, it is higher than 66.9% in Kenya [42], 63.1% in Zimbabwe, 72.0% in Oyo in Nigeria 72.0% [43] and 73.3% in Nigeria [44], Another study in Cameroon reported 62.6% [45] which are equally developing countries. The higher prevalence obtained in this study could be accounted for the fact that this study is conducted in a crises zone ‘‘Anglophone crises’’ which is a well-established perpetrator for mental health problems, financial stresses which are contributors to perpetration of family violence [2]Though social, cultural and economic factor difference play a key role in the prevalences, we believe other factors notably, anglophone crises, lockdowns crises related, including the additional COVID-19 pandemic measures and staying home without going to work increased time spent with partners and likelihood of violence or perception of it. Additionally, most of the study studies took into consideration just 3 forms of violence i.e. psychological, physical and sexual.

Psychological violence stood out as the most common form of violence experienced by pregnant women in this study. This study is in line with other studies that had psychological violence as most common form [18,19,47] but differ with other studies that reported sexual violence as most common form [46,21]. Notwithstanding, the prevalence of psychological violence is in line with that of Iran 88.4% [39] The difference in prevalence can be accounted as stated above from multiple stresses due to the anglophone crises and additional impact of COVID-19 with loss of jobs. The prevalence of physical violence according to this study was (42.2%) This value is within range of 33.1%-63.2% estimated by a systematic review conducted by Ashimi A and Amole T, 2015. [48]. Contrarily it is higher than figures gotten from studies around Asia of 3.2%-10.0% [46,15,42]. The observed difference may have been as a result of variation in cultures and lack of stresses during the time of study. The prevalence of sexual violence estimated in this study was 34.3%, This is similar to 33.7% obtained by Tsague, G. N. et al, 2024 in Yaounde Cameroon [50]. This might be accounted for by similarities in culture within the same culture. The prevalence of controlling behavior in our study was 78.0% which is lower than the prevalence gotten from a study by Esther et al in Nigeria with 98.8% [44] but far higher than that gotten from Ethiopia 30.4% [21]. The study analyzed life time prevalence while this study analyzed controlling behavior during index pregnancy. The prevalence of economic violence (42.2%) in our study is quite high compared to that seen in Ghana (7.4%0) and Ethiopia (27,0%) [21]. The difference between our study and that of Ghana could be due to the differences in study designs Ghana used a clustered randomize control trial. and social, economic and political differences and the period of data collection.

### Determinants of domestic violence in pregnancy

The results of this study reveal that pregnant women with partners who drank alcohol were more likely to be victims of domestic violence specifically psychological and controlling behavior violence. This finding is consistent with other findings in Ethiopia [13,20,21, 14], in Nigeria [44], and in Greece [15]. This association could be explained by the fact that alcohol consumption raises levels of aggression, misunderstanding of verbal or non-verbal actions and also alcohol consumption might be a source of dispute in relationships. Women who received less than 30,000 XAF a month were more than 3 times more likely of being exposed to psychological and controlling behavior violence. This study is in agreement with a study by Antoniou *et al* in Greece [15] and that of Akashi A. *et al* in Rwanda [16] and that of Abramsky T. *et al* in Tanzania [50]. This could occur because the women depend on the men for financial support, they are bound to endure all sorts of psychological and controlling behavior each time they go to ask for or need money. Whereas increasing income leads to less frequent argument, improved communication and increased women’s confidence and independence [50]. In this study, women who consume alcohol were 2.4 times more exposed to psychological violence, this is consistent with a systematic review carried out on studies in Africa [51] and a study in Brazil [35]. This could be explained by the fact that alcohol can increase impulsive behavior which can lead to violence. Notwithstanding this study did not access whether the alcohol use came before the violence or it was used as a coping strategy after experiencing violence. Women with lower than tertiary level of education were more at risk of physical violence the risk increased as the level of education reduces. Findings from prior studies had same reviews [18,52]. Also, studies conducted in Ethiopia found education be it lower or higher to be protective of violence [13,19]. This could be attributed to the fact that education increases the opportunities for employment, access to information on their rights and greater negotiating abilities and choice of partners as well as reducing dependency on their partners. Partners who were older than 40years were more likely to perpetrate physical violence. A study by Bikila *et a*l and Semahegn *et al* in Ethiopia also revealed that older partners were more likely to perpetrate violence [22,14]. whereas older partners were protective of sexual violence. The age discrepancy between the women and their partner could be the reason for the increased odds of violence amongst oldest intimate partners. Contrarily to physical violence, an older age partner was instead protective of sexual violence. Similar results were found in Nepal [46] and Zimbabwe [51]. The above can be explained by a possible liaison between a partner’s age increase and emotional growth as well as development of a social sense of responsibility thus leading to less inter spousal conflict. Partners who smoked had a higher probability of perpetrating physical and economic violence than those who did not smoke. This result was in accordance with prior studies in Gambia [53], in Ethiopia [21] and Iran [54]. In our study, being a housewife increased the chances for economic violence to occur. with the finding is similar to that of Fekadu, E, et al, 2018 in Ethiopia [55] this could be explained by the fact that housewives are more economic dependent on their partners which make them vulnerable and less autonomous which is more likely to cause disagreement and hence violence. On the contrary, a study carried out in Ethiopia, women who worked were more likely to experience violence [19]. Findings in this study also revealed that women who started ANC late were more than 2 times at risk of economic violence which is in concordance with a study in Ethiopia [21]. This may be due to the lower socio-economic status or lack of resources to access services and lack of knowledge on the advantage of on-time ANC initiation as they instead spend money on smoking and gambling.

This study relied on participants’ ability to recall past experiences of violence, which may have introduced recall bias, as some events might not have been accurately remembered. To mitigate this, a structured questionnaire with predefined response options was used to aid recall and enhance data reliability. Additionally, the study was conducted within a hospital setting, capturing domestic violence prevalence only among pregnant women ANC. As a result, the findings may not be fully generalizable to all pregnant women in the community, particularly those who do not attend ANC, who may be at an even higher risk of experiencing domestic violence.

#### Conclusion

Domestic violence during pregnancy is highly prevalent in the Buea Health District, with the majority of pregnant women experiencing at least one form of abuse. Psychological violence emerged as the most common type, primarily perpetrated by intimate partners. Several factors, including alcohol consumption, gambling, smoking, low education and income levels, large household sizes, and late initiation of antenatal care, were found to be associated with increased vulnerability to domestic violence. Conversely, women who had the autonomy to choose their partners were less likely to experience such abuse. These findings highlight the urgent need for targeted interventions to address domestic violence during pregnancy and support affected women.

## Data Availability

The dataset and materials of the study are available upon reasonable request from the corresponding author.

## Declarations

### Abbreviations

CI: Confidence Interval
DHS: Demographic Health Survey
LMICs: OR: Odds Ratio
SPSS: Statistical Package for the Social Science
WHO: World Health Organization

## Acknowledgments

We are grateful to the study participants and administrators of the health facilities for their collaboration in data collection.

## Authors’ Contributions

EAT supervised the study, participated in conception, literature review, design of the study, data analysis and revising the manuscript for publication. HNC participated in the conception, literature review, design of the study, data collection, interpretation and management, drafting, and revising the manuscript for publication. JAN supervised the study, participated in the conception, design of the study and review. SSN participated in the data analysis, validation, data interpretation and drafting of manuscript. MNL participated in literature review, data analysis, validation and drafting of manuscript. DSN participated in study conception, supervision and reviewed the manuscript for publication. All authors read and approved the final copy of manuscript.

